# Cardiovascular disease and subsequent risk of psychiatric disorders: a nationwide sibling-controlled study

**DOI:** 10.1101/2022.06.27.22276962

**Authors:** Qing Shen, Huan Song, Thor Aspelund, Jingru Yu, Donghao Lu, Jóhanna Jakobsdóttir, Jacob Bergstedt, Lu Yi, Patrick F. Sullivan, Arvid Sjölander, Weimin Ye, Katja Fall, Fang Fang, Unnur Valdimarsdóttir

## Abstract

**Background:** The association between cardiovascular disease (CVD) and selected psychiatric disorders has frequently been suggested while inadequate control of familial factors and comorbidities has halted causal inferences.

**Methods:** We identified 869 056 patients newly diagnosed with CVD from 1987 to 2016 in Sweden while no history of psychiatric disorders, and 910 178 full siblings of these patients and 10 individually age- and sex-matched unrelated population controls (N=8 690 560). Adjusting for multiple comorbid conditions, we used flexible parametric models and Cox regression to estimate the association of CVD with risk of all subsequent psychiatric disorders, comparing rates of first incident psychiatric disorder among CVD patients with rates among unaffected full siblings and population controls.

**Results:** The median age at diagnosis was 60 years for patients with cardiovascular disease and 59.2% were male. During up to thirty years of follow-up, the crude incidence rates of psychiatric disorder were 7.1, 4.6 and 4.0 per 1000 person-years for patients with cardiovascular disease, their siblings and population controls. In the sibling comparison, we observed an increased risk of psychiatric disorder during the first year after cardiovascular diagnosis (hazard ratio [HR], 2.74; 95% confidence interval [CI], 2.62-2.87) and thereafter (1.45; 95% CI, 1.42-1.48). Increased risks were observed for all types of psychiatric disorders and among all diagnoses of cardiovascular disease. We observed similar associations in the population comparison. Cardiovascular patients who developed a comorbid psychiatric disorder during the first year after diagnosis were at increased risk of subsequent cardiovascular death compared to patients without such comorbidity (HR 1.55; 95% CI 1.44-1.67).

**Conclusions:** Patients diagnosed with cardiovascular disease are at an elevated risk for subsequent psychiatric disorders independent of familial factors and comorbid conditions. Cardiovascular patients with comorbid psychiatric disorders are at increased risk of cardiovascular mortality suggesting that surveillance and treatment of psychiatric comorbidities should be considered as an integral part of clinical management of newly diagnosed patients with cardiovascular disease.

**Funding:** this work was supported by the EU Horizon 2020 Research and Innovation Action Grant (CoMorMent, Grant nr. 847776 to UV, PFS and FF), Grant of Excellence, Icelandic Research Fund (grant no. 163362-051 to UV), ERC Consolidator Grant (StressGene, grant no: 726413 to UV), Swedish Research Council (Vetenskapsrådet, award D0886501 to PFS) and US NIMH R01 MH123724 (to PFS).

## Introduction

Being diagnosed and living with a major life-threatening disease is stressful and associated with multiple biologic processes that combined may contribute to the development of psychiatric disorders. It is for instance firmly demonstrated that a cancer diagnosis is associated with a rapid rise of psychiatric disorders^1^ and self-inflicted injury,^2^ which further is associated with a compromised cancer specific survival.^3^ Psychiatric comorbidities have also been reported among patients with cardiovascular disease (CVD), e.g. after stroke,^4^ heart failure,^5^ and myocardial infarction.^6,7^ with indications of elevated risk of overall mortality.^8,9^ Yet, evidence on the association between CVD and subsequent development of psychiatric disorders is still limited as previous research has mainly relied on selected patient populations instead of complete follow-up of general population as well as limited control of reverse causality and important confounding factors, e.g. familial factors and comorbidities.^10–12^

We thoroughly searched the existing literature of investigating the association between CVD and clinically confirmed psychiatric disorders or psychiatric symptoms. After excluding most studies with either cross-sectional or retrospective designs, we only found 12 prospective cohort studies investigating the risk of selected psychiatric disorders following diagnosis of cardiovascular disease (summarized in Table S1, Supplementary Appendix). While these prospective studies suggest a positive association between hypertension, heart disease, and stroke with depressive symptoms^11,13–15^ and stress-related disorders,^16^ they were limited to elderly populations,^17,18^ and using self-reported ascertainment of psychiatric outcomes.^17,18^ Only few of these studies addressed incident or first diagnosed psychiatric disorders among patients with CVD, e.g. exlcuding patients with history of psychiatric disorders, and no study addressed the issue of familial confounding. Indeed, genetic correlation between these two complex diseases have recently been documented ^19–21^ as well as the importance of early life environment for the development of both cardiovascular disease^22^ and psychiatric disorders.^23^ It is therefore unknown whether the reported association between CVD and psychiatric disorder can be attributed to unmeasured confounding shared within families.^19–21^ Thus, a comprehensive evaluation of the association between all CVDs and risk of any incident psychiatric disorder, addressing the abovementioned shortcomings, is warranted.

With up to thirty years of follow-up and with nationwide complete information on family links in Sweden, we aimed to comprehensively investigate the association between CVD diagnosed in specialist care and subsequent risk of incident psychiatric disorders while accounting for familial factors through a sibling comparison. We further aimed to estimate the potential role of psychiatric comorbidity in cardiovascular mortality among patients with CVD.

## Materials and Methods

### Study Design

The Swedish Patient Register contains national information on inpatient care with complete coverage since 1987 and outpatient specialized care since 2001.^25^ The Swedish Multi-Generation Register includes nearly complete familial information for Swedish residents born since 1932.^26^ Using personal identification numbers assigned to all Swedish residents, we identified all individuals born in Sweden after 1932 who received a first diagnosis of any CVD that attended in inpatient or outpatient specialized care between 1 January 1987 and 31 December 2016 (N=986 726). Patients diagnosed with CVD before age 5 (N=6 091, probable congenital heart disease) or with a history of any psychiatric disorder before the diagnosis of CVD (N=111 579) were excluded, leaving 869 056 patients in the analysis (Fig S1, Supplementary Appendix). Date of first CVD diagnosis was used as the index date for the exposed patients.

Through the Multi-Generation Register, we identified all full siblings of patients with CVD (58.6% of all CVD patients) who were alive and free of CVD and psychiatric disorder at the time when their affected sibling was diagnosed (N=910 178). In addition, for each patient with CVD, we randomly selected 10 age-and sex-matched individuals from the general population who were free of CVD or psychiatric disorder when the index patient was diagnosed (N=8 690 560). The date of CVD diagnosis for the index patient was used as the index date for their unaffected siblings and matched population controls.

All study participants were followed from the index date until first diagnosis of any psychiatric disorder, death, emigration, first diagnosis of CVD (for unaffected siblings and matched population controls), or the end of the study period (31 December 2016), whichever occurred first.

### Ascertainment of Cardiovascular Disease and Psychiatric Disorder

We defined CVD as any first inpatient or outpatient hospital visit with CVD as the primary diagnosis from the Swedish Patient Register. Incident psychiatric disorder was defined as any first inpatient or outpatient hospital visit with psychiatric disorder as the primary diagnosis. We used the 9^th^ and 10^th^ Swedish revisions of the International Classification of Diseases (ICD-9 and 10) codes to identify CVD and psychiatric disorders and their subtypes (Table S2, Supplementary Appendix). In line with previous study,^27^ we classified CVD as ischemic heart disease, cerebrovascular disease, emboli/thrombosis, hypertensive disease, heart failure, and arrhythmia/conduction disorder. We classified psychiatric disorders as non-affective psychotic disorders, affective psychotic disorders, alcohol or drug misuse, mood disorders excluding psychotic symptoms, anxiety and stress-related disorders, eating disorders, and personality disorders.^28^

### Covariates

We extracted socioeconomic information for each participant, including educational level, individualized family income and cohabitation status, from the Longitudinal Integration Database for Health Insurance and Labor Market.^29^ Missing information on socioeconomic status was categorized as unknown or missing group. A history of somatic diseases was defined as having any of the following conditions before the index date: chronic pulmonary disease, connective tissue disease, diabetes, renal diseases, liver disease, ulcer diseases, malignancies, and HIV infection/AIDS (Table S2, Supplementary Appendix). We defined a family history of psychiatric disorders as a diagnosis of any psychiatric disorder among biological parents and full siblings of the study participants before the index date according to the Swedish Patient Register.

### Statistical Analysis

We used flexible parametric survival models to estimate the time-varying association between CVD and subsequent risk of incident psychiatric disorders,^30^ by comparing the rates of incident psychiatric disorders in CVD patients with the corresponding rates in their unaffected full siblings and matched population controls. As we observed a marked risk increase of psychiatric disorders immediately following the CVD diagnosis, we separately assessed the association within one year of CVD diagnosis and beyond one year. Hazard ratios (HRs) and their 95% confidence intervals (CIs) were derived from stratified Cox regression models, using time since the index date as the underlying time scale. We estimated HRs for any psychiatric disorder and categories of psychiatric disorders. We performed subgroup analyses by sex, age at index date (<50, 50-60, or >60 years), age at follow-up (<60 or ≥60 years), calendar year at index date (1987-1996, 1997-2006, or 2007-2016), history of somatic diseases (no or yes), and family history of psychiatric disorder (no or yes). In the sibling comparison, all Cox models were stratified by sibling sets and adjusted for sex, birth year, educational level, individualized family income, cohabitation status, and history of somatic diseases. In the population comparison, all Cox models were stratified by the matching variables birth year and sex and adjusted for all abovementioned covariates plus family history of psychiatric disorder.

To study the impact of additional cardiovascular comorbidity (i.e., patients with another type of CVD after diagnosis of the first CVD), we analyzed the association by presence or absence of cardiovascular comorbidity after the index date according to the type of first CVD. This analysis was restricted to follow-up beyond one year to focus on patients who survived their first CVD to be able to receive the diagnosis of another CVD. As a patient with CVD might have different types of CVD, we identified all diagnoses of CVD during follow-up and considered CVD comorbidity as a time-varying variable through splitting the person-time according to each diagnosis.

Because the Swedish Patient Register includes only information related to specialist care, we might have misclassified patients with psychiatric symptoms who were only attended by primary care. In a sensitivity analysis, we excluded also individuals with prescribed use of psychotropic drugs before the index date (ascertained from the Swedish Prescribed Drug Register including information on all prescribed medication use in Sweden since July 2005), and followed the remaining participants during 2006-2016. Use of psychotropic drugs during follow-up was also considered as having psychiatric disorder in this analysis.

To study rate of cardiovascular mortality (ascertained from the Swedish Causes of Death Register) in relation to psychiatric comorbidities after CVD diagnosis, we plotted Kaplan-Meier survival curves beyond the first year of follow-up for CVD patients with or without a diagnosis of psychiatric disorder during the first year of follow-up, separately. In addition to this 1-year time window, we also studied six months or two years since CVD diagnosis, to assess the robustness of these survival curves. We calculated the HRs of cardiovascular mortality for these two groups of patients using Cox regression model.

Analyses were performed in STATA 17.0 (StataCorp LP). All tests were two sided and P<0.05 was used as statistically level of significance. The study was approved by the Ethical Vetting Board in Stockholm, Sweden (DNRs 2012/1814-31/4 and 2015/1062-32).

## Results

The median age at index date was 60 years for CVD patients and 55 years for their unaffected full siblings (Table 1). 59.2% of the CVD patients and 48.4% of their unaffected siblings were male. CVD patients were more likely to have a history of somatic diseases than their unaffected siblings and matched population controls (15.6% vs. 8.8% and 11.0%). The most common diagnoses among the CVD patients were ischemic heart diseases (24.5%), arrhythmia/conduction disorders (24.2%), and hypertensive diseases (17.3%). The majority of the CVD patients had only one CVD diagnosis (without additional CVD comorbidities) during follow-up (69.7%).

**Table 1.**
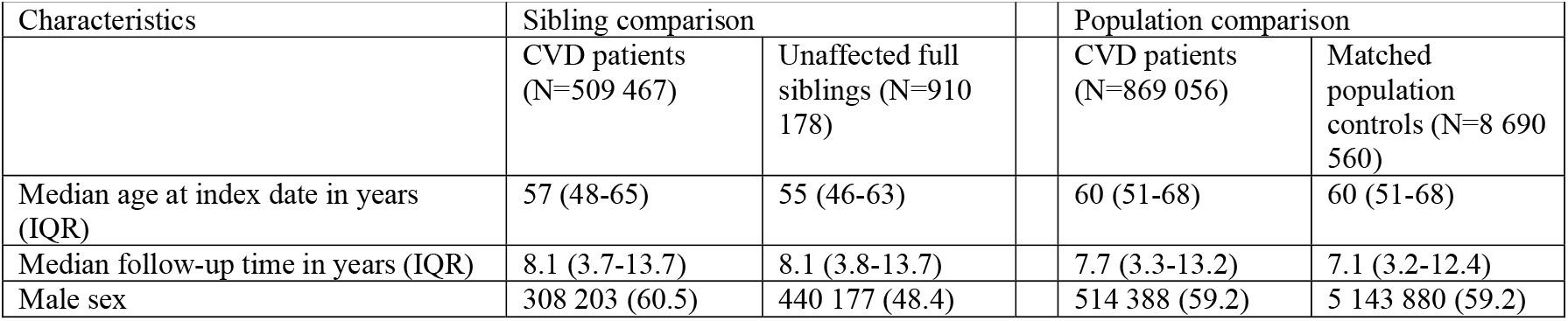
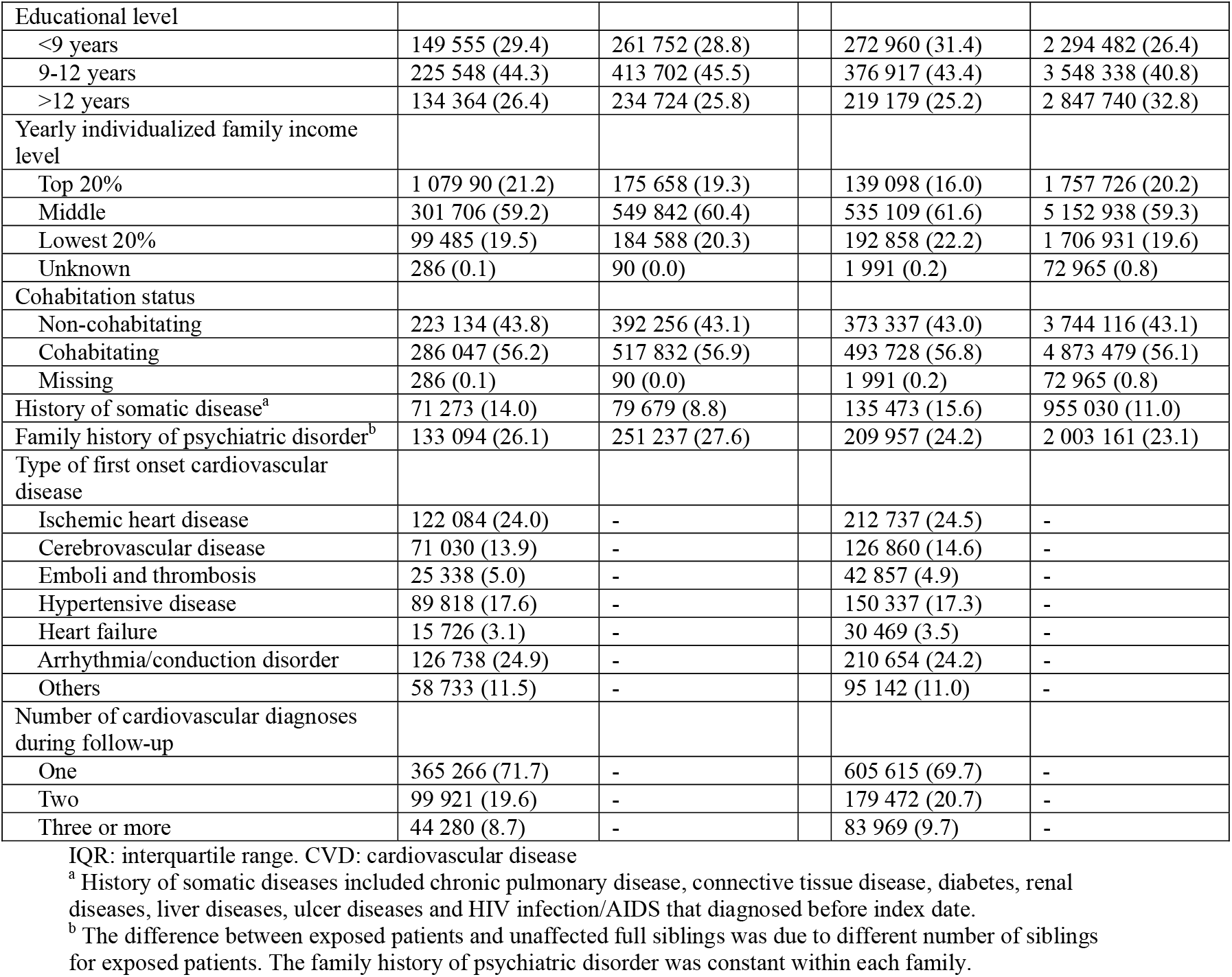
Characteristics of CVD patients diagnosed in Sweden between 1987 and 2016, their unaffected siblings and matched population controls.

During up to thirty years of follow-up, the crude incidence rates of psychiatric disorder were 7.1, 4.6 and 4.0 per 1000 person-years among CVD patients, their unaffected full siblings, and matched population controls, respectively (Table S3, Supplementary Appendix). Compared with unaffected siblings, CVD patients showed an increased risk of incident psychiatric disorder, especially immediately after diagnosis (Fig 1). The risk declined rapidly within the first few months after diagnosis and decreased gradually thereafter: the HR was 2.74 (95% CI 2.62 to 2.87) within first year and 1.45 (95% CI 1.42 to 1.48) beyond first year (Table S4, Supplementary Appendix). The risk increment was noted in all types of psychiatric disorders within and beyond first year of follow-up (Fig 2). Overall, the observed risk increase was similar in sibling and population comparisons, although the HR of non-affective psychotic disorders beyond one year of CVD diagnosis was smaller in the sibling comparison than in the population comparison. During the entire follow-up, we found similar positive associations across sex, age at index date, age at follow-up, history of somatic diseases and family history of psychiatric disorders (Table S3). A greater risk increment was observed in recent calendar years than earlier years.

**Figure 1.**
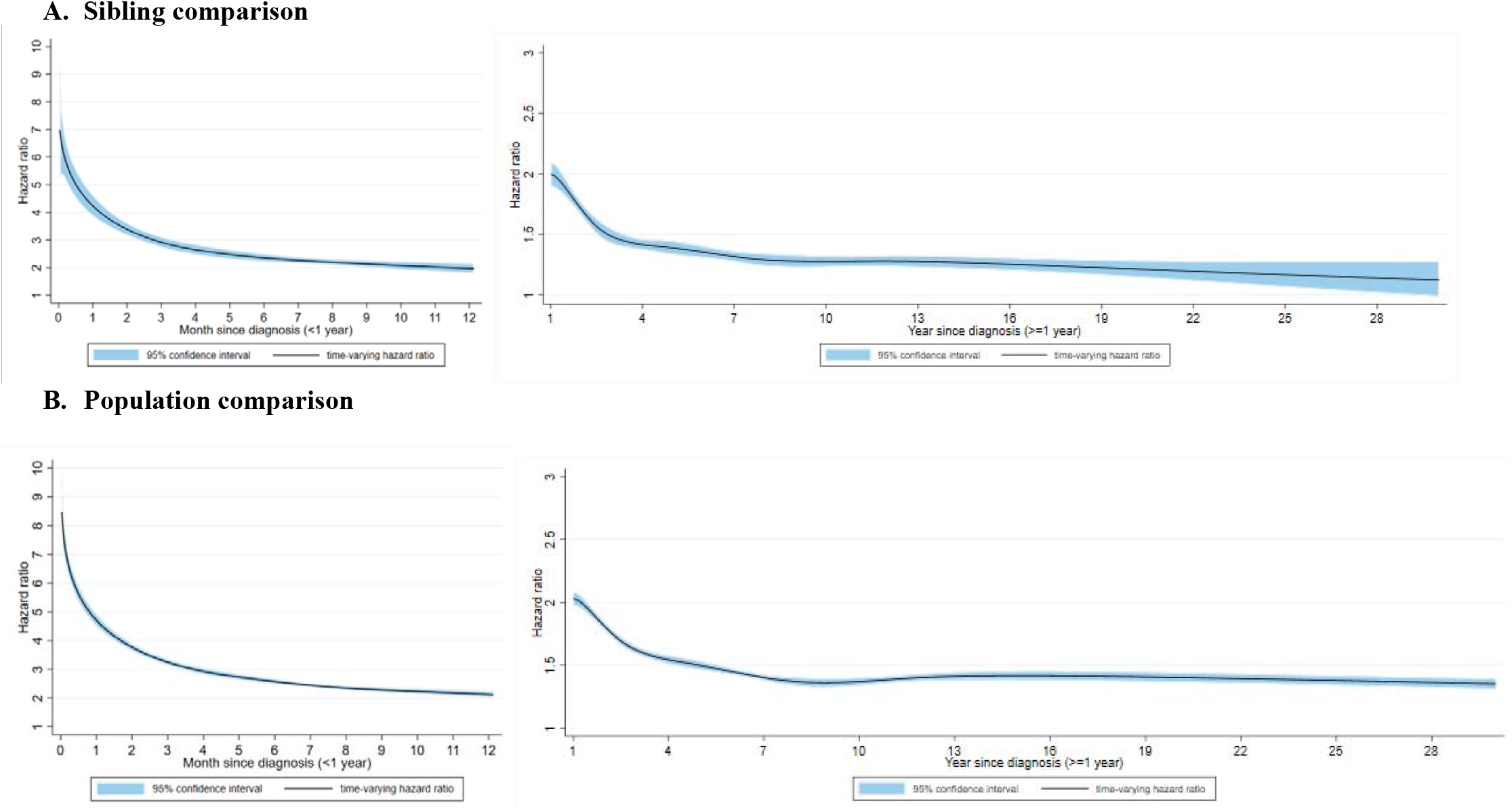
Time-varying hazard ratios for an incident psychiatric disorder among CVD patients, compared with their unaffected full siblings (sibling comparison) or matched population controls (population comparison), by time of follow-up (<1 year and >=1 year from CVD diagnosis)^*^ * CVD: cardiovascular disease. Time-varying hazard ratios and 95% confidence intervals were derived from flexible parametric survival models, allowing the effect of psychiatric disorder to vary over time. A spline with 5 df was used for the baseline rate, and 3 df was used for the time-varying effect. All models were adjusted for age at index date, sex, educational level, yearly individualized family income, cohabitation status, history of somatic diseases, as well as family history of psychiatric disorder (for population comparison).

**Figure 2.**
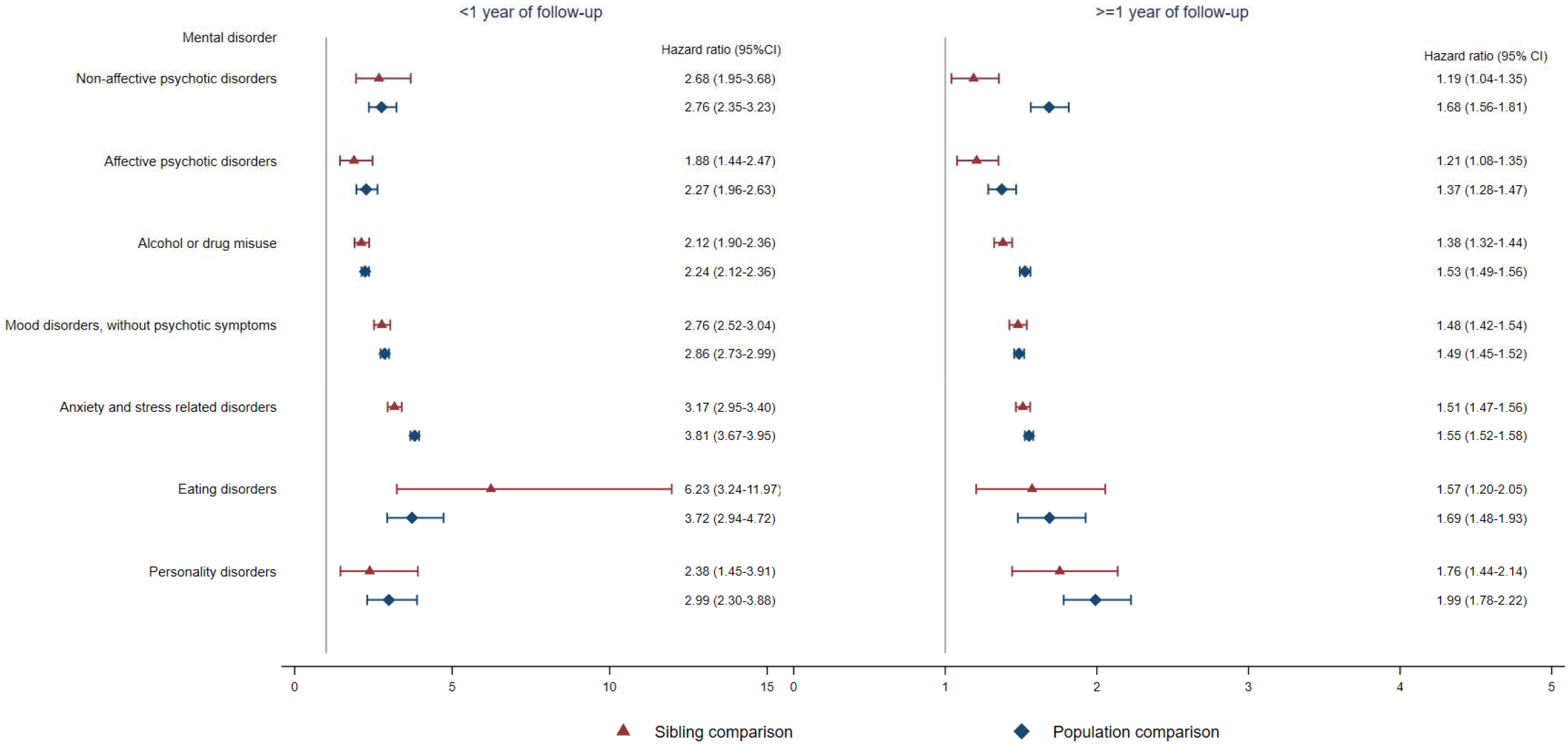
Hazard ratios with 95% confidence intervals for different types of psychiatric disorder among CVD patients compared with their full siblings and matched population controls, by time of follow-up (<1 or >=1 year from CVD diagnosis)^*^ * CVD: cardiovascular disease. Cox regression models were stratified by family identifier for sibling comparison or matching identifier (birth year and sex) for population comparison, controlling for age at index date, sex, educational level, individualized family income, cohabitation status, history of somatic diseases, and family history of psychiatric disorder (in population comparison). Time since index date was used as underlying time scale.

We found an increased risk of incident psychiatric disorder among all groups of CVD patients, with the most marked risk increase observed among patients with cerebrovascular disease and heart failure (Fig 3). A greater risk increase of incident psychiatric disorder was noted among CVD patients with additional cardiovascular comorbidities beyond one year of first CVD diagnosis compared with CVD patients without such comorbidities, except among those with heart failure (Fig S2, Supplementary Appendix). We found a similar positive association between CVD and risk of incident psychiatric disorder, when including use of psychotropic drugs as a definition of psychiatric disorder (Table S5, Supplementary Appendix).

**Figure 3.**
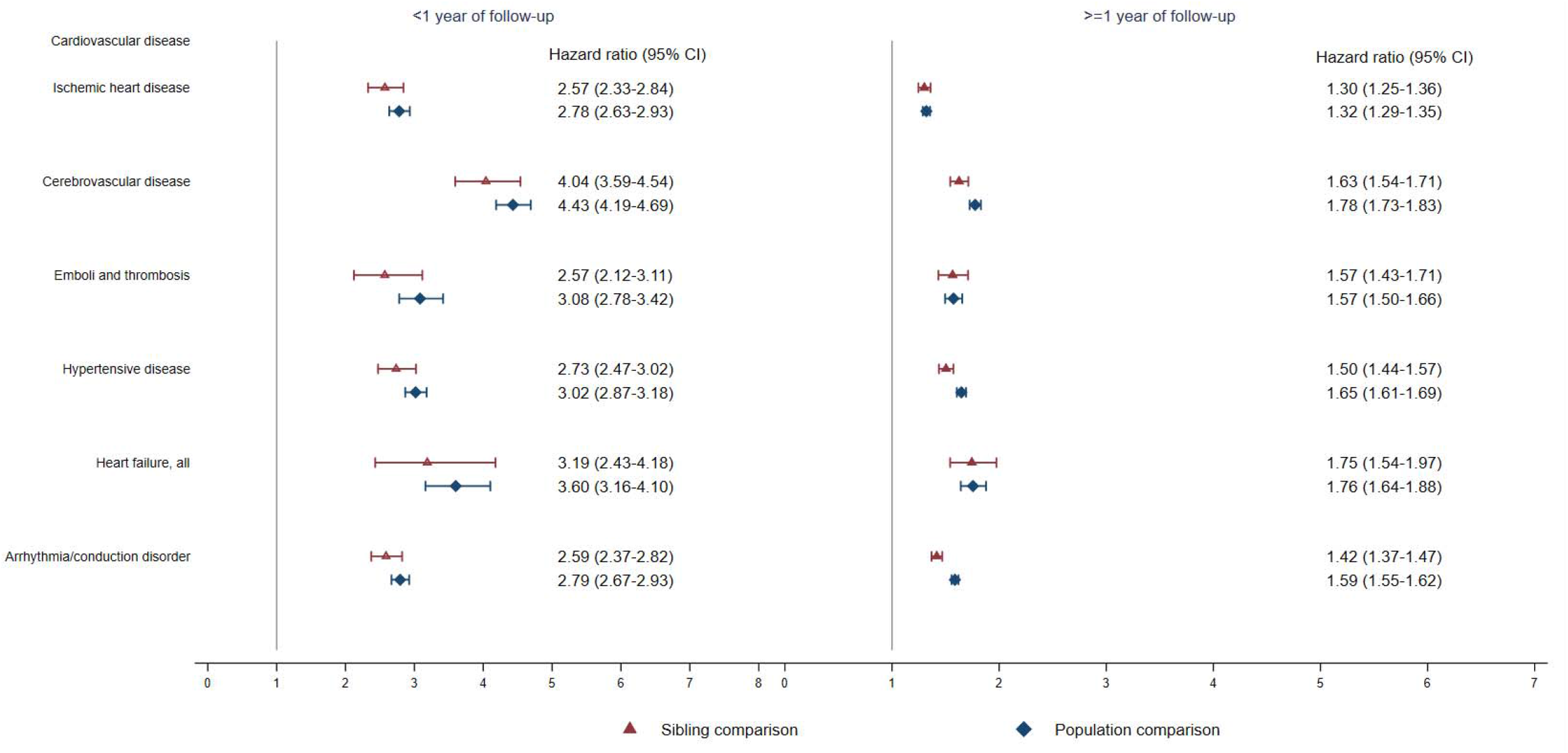
Hazard ratios with 95% confidence intervals for psychiatric disorders among different groups of CVD patients compared with their full siblings and matched population controls, by time of follow-up (<1 or >=1 year from CVD diagnosis)^*^ * CVD: cardiovascular disease. Cox regression models were stratified by family identifier for sibling comparison or matching identifier (birth year and sex) for population comparison, controlling for age at index date, sex, education level, individualized family income, cohabitation status, history of somatic diseases, and family history of psychiatric disorder (in population comparison). Time since index date was used as underlying time scale. We identified all cardiovascular diagnoses during follow-up and considered CVD comorbidity as a time-varying variable by grouping the person-time according to each diagnosis.

CVD patients diagnosed with subsequent psychiatric disorder showed a reduced CVD-specific survival compared with patients without such diagnosis (Fig 4, and Supplementary Appendix Fig S3 and S4). The HR of cardiovascular death was 1.55 (95% CI 1.44 to 1.67) when comparing CVD patients with a diagnosis of psychiatric disorder to patients without such diagnosis (mortality rate, 9.2 and 7.1 per 1 000 person-years, respectively).

**Figure 4.**
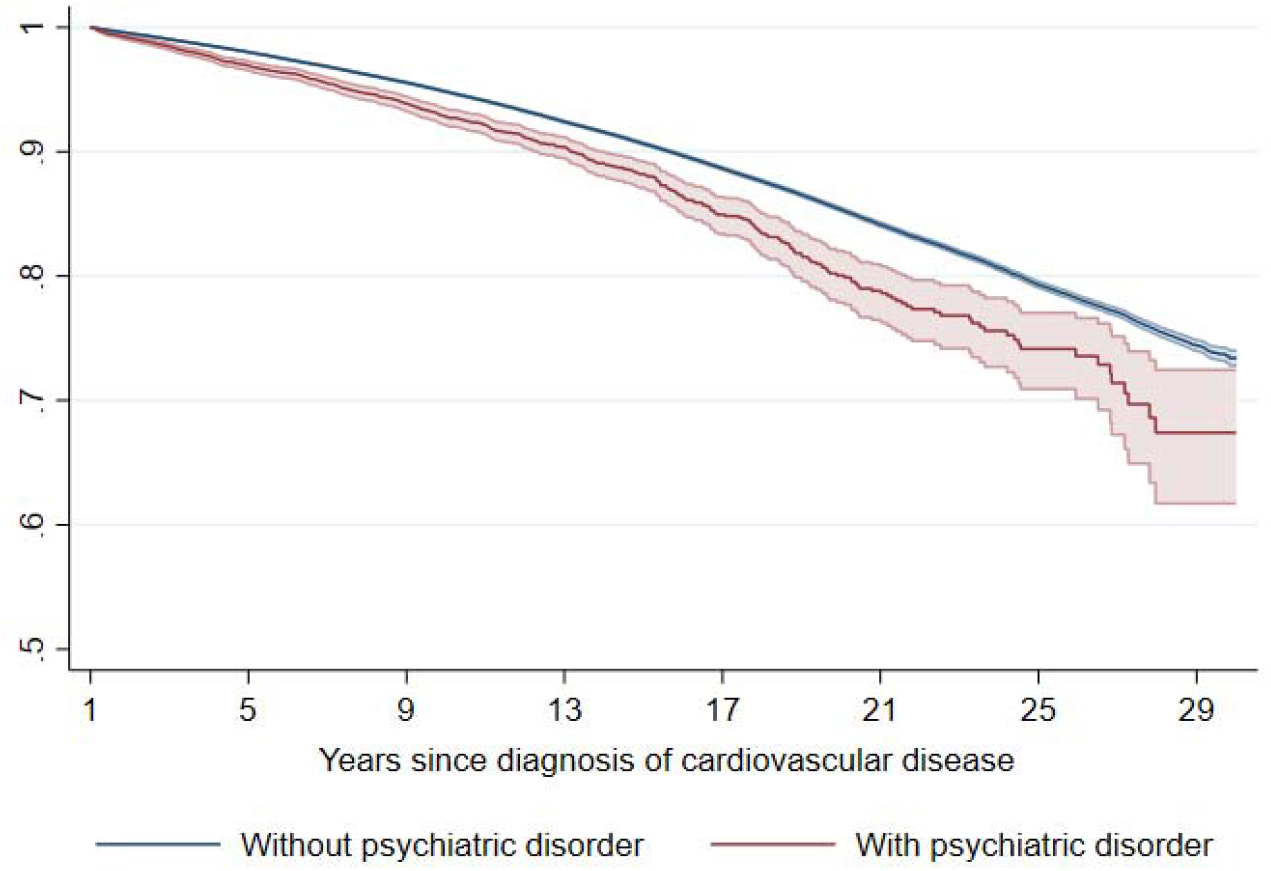
Kaplan-Meier estimates of CVD death in CVD patients with and without incident psychiatric disorder during the first year of follow-up ^*^. ^*^ CVD: cardiovascular disease. Time since index date was used as underlying time scale. 90.4% of CVD patients (N=785 287) survived the first year of follow-up and included in this analysis.

## Discussion

Our large population-based sibling-controlled cohort study including all patients diagnosed with their first CVD between 1987 and 2016 in Sweden, their unaffected full siblings, as well as a set of randomly selected unaffected population controls reveals a robust association between CVD and subsequent risk of incident psychiatric disorder. We found that patients with CVD were at elevated risk of various types of psychiatric disorders, independent of confounding factors shared within families and history of somatic diseases. The risk increment was greatest during the year after CVD diagnosis, indicating an opportunity for clinical surveillance in a high-risk time window. Further, an occurrence of psychiatric comorbidity after CVD diagnosis was associated with an approximately 55% increased risk of subsequent death from cardiovascular causes. This finding further underscores the importance of surveillance and, if needed, treatment of psychiatric comorbidities among newly diagnosed CVD patients.

### Strengths and Limitations

The strengths of our study include its large sample size of the entire Swedish nation and the prospective study design with sibling comparison that significantly alleviates concerns of familial confounding from shared genetic and early-life environmental factors between siblings. The Swedish population and health registers provide the opportunity to obtain complete follow-up as well as prospectively and independently collected information on disease identification, minimizing the risk of selection and information biases. The large sample size of our study further enables detailed subgroup analyses by types of CVD, types of psychiatric disorders, and patient characteristics.

Some limitations need to be acknowledged. First, we identified patients with CVD and psychiatric disorder through inpatient or outpatient hospital visit. The later inclusion of outpatient records in the Swedish Patient Register may lead to underestimation of the actual numbers of patients with CVD and psychiatric disorder, especially those with relatively milder symptoms. Second, we missed individuals attended in primary care, which may underestimate the proportion of individuals with history of psychiatric problems at study entry. To alleviate such concerns, we additionally considered the use of prescribed psychotropic drugs and found similar results. Further, patients with CVD have an established contact with health care and may therefore be more likely than others to be diagnosed with psychiatric disorder. Although such surveillance bias may to some extent explain the increased risks during the first few months after CVD diagnosis, it is unlikely that the risk elevation during the entire follow-up is attributed to such bias. Finally, we could not directly control for lifestyle factors (e.g., smoking, diet, and physical exercise) and therefore cannot exclude the possibility of residual confounding not fully controlled for in the sibling comparison.

### Comparison with other studies

Our findings are consistent with the existing literature suggesting a positive association between CVD and different types of psychiatric disorders. In previous studies, an increased risk of depression^13–15^ and anxiety^11^ was noted after diagnosis of stroke,^11,13–15^ hypertension,^17^ coronary artery disease^31^ and atrial fibrillation.^32^ Such risk elevations has been suggested to be persistent over time,^10,33,34^ and, in parallel with our findings, associated with compromised survival.^8,35,36^ However, the evidence from prospective cohort studies with a long and complete follow-up as well as with a thorough control of confounding factors and comorbid conditions has, up to this point, been limited. Our study therefore complements previous findings revealing a positive association between a broader range of CVDs and subsequent risk of incident psychiatric disorder, using a large cohort with control of various confounding factors. We found the association to be robust both in sibling and population comparisons, and after additional adjustment for various comorbidities including other additional CVDs, indicating that the association is unlikely explained by shared familial factors and various comorbidities. We showed that CVD patients with psychiatric comorbidity was associated with an increased risk of subsequent CVD death, highlighting the importance of surveillance and prevention on psychiatric comorbidities for those newly diagnosed CVD patients.

The association between CVD and psychiatric disorders was noted both in men and women, across all age groups and calendar periods, as well as among individuals with or without a history of somatic diseases and family history of psychiatric disorders. Previous studies have indicated that major depression was more commonly recognized among individuals with multimorbidity (more than one CVD diagnosis) than those with only one condition.^37^ In our study, about 30% of the CVD patients developed one or more cardiovascular comorbidities during follow-up, and a higher risk of psychiatric disorder was indeed noted among patients with multiple CVD diagnoses. A compromised survival from CVD cause was indeed observed among most patients with common types of first CVD diagnosis and comorbid with psychiatric disorder.

### Potential mechanisms

The pathophysiological mechanisms linking CVD and psychiatric disorders are complex and not well understood, and may vary with specific diagnoses of CVD and psychiatric disorders. The highly increased risk noted immediately after CVD diagnosis may indicate a direct impact of stress reaction of being diagnosed with a life-threatening disease.^38^ In addition, it has been proposed that biological alterations in the cardiovascular system to a severe stress response may increase the risk of various psychiatric disorders.^39^ For example, cardiovascular risk factors including hypercoagulability, dyslipidemia, and an impaired immune response have been associated with impaired psychological health.^39^ Some biological changes in patients with coronary heart disease (e.g., decreased heart rate variability, increased arterial stiffness, and endothelial dysfunction) have been observed in patients with depressive and anxiety disorders.^40–42^ Chronic inflammation may induce the development of atherosclerosis and arterial thrombosis, and elevation in inflammatory biomarkers (i.e., IL-6 and C-reactive protein) has been reported in various psychiatric disorders including post-traumatic stress disorder and major depression.^43–45^ Other behavioral and psychosocial factors may as well interact with these pathways and need to be understood further.

## Conclusions

Using a large population-based sibling-controlled cohort with up to thirty years of follow-up, we demonstrate that patients with CVD have an increased risk of first-diagnosed psychiatric disorder, independent of familial background, history of somatic diseases, and other cardiovascular comorbidities. Our study further indicates an impact of psychiatric comorbidities on cardiovascular mortality among patients with CVD, motivating increased surveillance of psychiatric comorbidity among newly diagnosed patients with CVD.

## Data Availability

Data analyses were performed in STATA 17.0 (StataCorp LP). STATA script used in the primary analyses has been made available as supplementary appendix. Aggregated data used for generating figures are available in supplementary appendix. The original data used in this study are owned by the Swedish National Board of Health and Welfare and Statistics Sweden. The authors are not able to make the dataset publicly available according to the Public Access to Information and Secrecy Act in Sweden. Any researchers (including international researchers) interested in accessing the data can send request to the authorities for data application by: 1) apply for ethical approval from local ethical review board; 2) contact the Swedish National Board of Health and Welfare (https://bestalladata.socialstyrelsen.se/) and/or Statistics Sweden (https://www.scb.se/vara-tjanster/bestall-data-och-statistik/) with the ethical approval and submit a formal application for access to register data. The same contacts can be used for detailed information about how to apply for access to register data for research purposes."

## Author Contributions

Drs Shen, Fang, and Valdimarsdóttir had full access to all the data in the study and take responsibility for the integrity of the data and the accuracy of the data.

*Concept and design*: Shen, Fang, and Valdimarsdóttir.

*Acquisition, analysis, or interpretation of data*: All authors.

*Drafting of the manuscript*: Shen, Fang, and Valdimarsdóttir.

*Statistical analysis*: Shen, Song, Fang, and Valdimarsdóttir.

*Obtained funding*: Fang, Valdimarsdóttir and Sullivan.

*Administrative, technical, or material support*: Shen, Fang, Valdimarsdóttir, Yu and Ye.

*Supervision*: Fang and Valdimarsdóttir.

## Ethical approval

The study was approved by the Ethical Vetting Board in Stockholm, Sweden (DNRs 2012/1814-31/4 and 2015/1062-32). Informed consent to each participant was waived by Swedish law in nationwide registry data.

## Transparency declaration

UV affirms that the manuscript is an honest, accurate, and transparent account of the study being reported; that no important aspects of the study have been omitted; and that any discrepancies from the study as planned have been explained.

## Supplementary Appendix

**Table S1.** Summary of prospective cohort studies addressing the association between various indications of cardiovascular disease and risk of psychitric disorders/psychiatric symptoms.

| Paper | Study design and database | Exposure | Outcome | Sample size and follow-up | Main findings | Control for |  |
| --- | --- | --- | --- | --- | --- | --- | --- |
|  |  |  |  |  |  | familial factors? | other cardiovascular comorbidities? |
| Petersson S, 2014<sup>1</sup> | population-based cohort study; baseline measured between 2000 and 2006 | hypertansion and stroke in structured interviews | incident depressive disorders | 567 very old people≥85 years; with 5 years of follow-up; . | hypertension OR 2.83 (1.08-7.42) and history of stroke OR 3.25 (1.12-9.44) | no | no |
| Zhang X, 2015<sup>2</sup> | Community-based study; random sampling between March 1999 and February 2001 | arrhythmia and heart failgure in self-report questionnaire | Generalized anxiety disorder (GAD) | 1711 individuals aged 65 years and above; median follow-up time was 9.7 years | arrhythmia and heart failure HR 1,67 (1.04-2.70) | no | no |
| Chang JC, 2017<sup>3</sup> | longidtudinal cohort study; a commuity health-screening program between 1999-2004 in Taiwan | clinical diagnosis of cardiovascular disease | clinical diagnosis of incident posttraumatic stress disorder (PTSD) | 76,417 aged 30 years and above and mean follow-up of 3.13 (1-5) years | cardiovascular disease HR 1.45 (1.03–2.04) | no | no |
| Feng HP, 2016<sup>4</sup> | cohort study; National Health Insurance Research Database in Taiwan during January 1 and December 31, 2005 | clinically diagnosed myocardial infarction | clinical diagnosis of anxiety or depressive disorder | 1396 patients with myocardial infarction and matched 13,960 individuals without MI (63% men); follow-up of 2 years | HR 5.06 (4.61–5.54) for anxiety disorders and HR 7.23 (4.88–10.88) for depressive disorders | no | control for hypertension and cerebrovascular disease in model |
| Kivimäki M, 2012<sup>5</sup> | prospective cohort study; London-based office staff, self-reported questionnaire and medical records | Vascular risk assessed with the Framingham Cardiovascular, Coronary Heart Disease, and Stroke Risk Scores | New depressive symptoms identified by General Health Questionnaire (GHQ) $\geq 5$, and Center for Epidemiologic Studies Depression Scale score (CES-D) $\geq 16$, and use of antidepressant medication. | 5318 participants (mean age 54.8 years, 31% women); baseline examination in 1991 and follow-up screenings in 1997, 2003, and 2008 | Diagnosed vascular disease (coronary heart disease or stroke) with an increased risk for depressive symptoms, OR 1.5 (1.0–2.2) to 2.0 (1.4–3.0) depending on indicators | no | no |
| Zimmerman JA, 2009<sup>6</sup> | Logitudinal study among community dwelling elder Mexican Americans during 1993-1994 | self-reported vascular risk factors (hypertension, stroke and myocardial infarction) | Depressive symptom Measured with Center for Epidemiologic Studies Depression Scale (CES-D $\geq 16$) | 964 participants (mean age 72) with two year of follow-up; 53% women | Hypertension: OR 2.3 (1.39-3.70); stroke: OR 0.83 (0.25-2.74); myocardial infarction: 1.48 (0.71-3.10) | no | no |
| Kim JM, 2006<sup>7</sup> | community-based prospective study from 2001-2003 | self-reported vascular disease risk (stroke, heart disease, hypertension) | Depression assessed using the community version of the Geriatric Mental State (GMS) | 661 community participants (mean age 73) with two year of follow-up; 55% women | Pre-existing heart disease: OR 2.2 (1.3-3.7); incident stroke: OR 2.2 (1.0-5.0) | no | no |
| Mast BT, 2008<sup>8</sup> | Community residents | prevalent vascular disease | incidence depressive symptoms using Center for Epidemiologic Studies Depression (CES-D $> 8$) scale. | 1,796 elders ages 70–79 with two year of follow-up; | Coronary heart disease: OR 1.68 (1.09-2.59); myocardial infarction: OR 2.69 (0.99-7.28); atrial fibrillation: OR 0.80 (0.19-3.42); left ventricular hypertrophy: OR 1.17 (0.65-2.12); congestive heart failure: OR 0.90 (0.27-3.06); hypertension: OR 1.07 (0.73-1.57); peripheral arterial disease: OR 1.58 (0.75-3.33) | no | Charlson index |
| Godin O, 2012<sup>9</sup> | multicenter cohort study with random sample during 1999-2010 | Measures of systolic and diastolic blood pressure or using antihypertensive medication | major depressive symptoms according to the Mini-International Neuropsychiatric Interview or taking antidepressant medication | 3,090 with ten years of follow-up; 59.9% female | Hypertension: RR 0.97 (0.76-1.22) | no | no |
| Garcia-Fabela L, 2009<sup>10</sup> | longitudinal study of community-dwelling persons | self-reported hypertension | depressive symptoms | 3,276 persons aged 60 years and older with two year of follow-up; | Hypertension: OR 1.18 (1.01–1.40) | no | Adjust for stroke, ischemic cardiopathy in model |
| Simning A, 2018<sup>11</sup> | nationally representative longitudinal survey of U.S. Medicare beneficiaries initiated in 2011 | new onset heart attack or stroke in interview | PHQ-2 identified clinically significant depressive symptoms | 5,643 aged 65 years and older with one year follow-up | Among those no close contact: heart attack OR 5.57 (1.68-18.44), stroke OR 4.44 (1.03-19.14); among those with close contacts: heart attack OR 1.19 (0.68-2.07), stroke OR 2.51 (1.76-3.59) | no | Adjust for medical conditions (include hypertension and heart disease) |
| Chuang CS, 2014<sup>12</sup> | Cohort study using National Health Insurance (NHI) data sets from Taiwan in period 2000-2002 | hyperlipidemia (comorbidity as hypertension) from insurance database | an insurance claim for diagnosis and treatment of depression | 26,852 exposed and 107,408 unexposed individuals | Hypertension: HR 1.70 (1.57–1.83) | no | no |

<sup>1</sup>Petersson, S. et al (2014). “Risk factors for depressive disorders in very old age: a population-based cohort study with a 5-year follow-up.” Soc Psychiatry Psychiatr Epidemiol 49, 831–839
<sup>2</sup> Zhang, X. et al (2015). “Risk factors for late-onset generalized anxiety disorder: results from a 12-year prospective cohort (The ESPRIT study).” Transl Psychiatry 5, e536
<sup>3</sup> Chang, JC. et al (2017). “Comorbid diseases as risk factors for incident posttraumatic stress disorder (PTSD) in a large community cohort (KCIS no.PSY4).” Sci Rep 7, 41276.
<sup>4</sup> Feng, H-P et al. “Risk of anxiety and depressive disorders in patients with myocardial infarction: A nationwide population-based cohort study.” Medicine vol. 95,34 (2016): e4464.
<sup>5</sup> Kivimäki M et al (2012). “Vascular risk status as a predictor of later-life depressive symptoms: a cohort study.” Biological Psychiatry;72:324 –330
<sup>6</sup> Zimmerman JA et al (2009). “Vascular risk and depression in the Hispanic Established Population for the Epidemiologic Study of the Elderly (PESE).” Int J Geriatr Psychiatry; 24: 409–416.
<sup>7</sup> Kim JM et al (2006). “Vascular risk factors and incident late-life depression in a Korean population.” Br J Psychiatry; 189: 26–30.
<sup>8</sup> Mast BT et al (2008). “Vascular disease and future risk of depressive symptomatology in older adults: findings from the Health, Aging, and Body Composition study.” Biol Psychiatry; 64: 320–326.
<sup>9</sup> Godin O et al (2012). “Body mass index, blood pressure, and risk of depression in the elderly: a marginal structural model.” Am J Epidemiol; 176: 204–213.
<sup>10</sup> Garcia-Fabela L et al (2009). “Hypertension as a risk factor for developing depressive symptoms among community-dwelling elders.” Rev Invest Clin; 61: 274–280.
<sup>11</sup> Simning A et al (2018). “The association of a heart attack or stroke with depressive symptoms stratified by the presence of a close social contact: findings from the National Health and Aging Trends Study Cohort.” International journal of geriatric psychiatry vol. 33,1: 96-103.
<sup>12</sup> Chuang CS et al (2014). “Hyperlipidemia, statin use and the risk of developing depression: a nationwide retrospective cohort study.” Gen Hosp Psychiatry;36(5):497-501.

**Figure S1.**
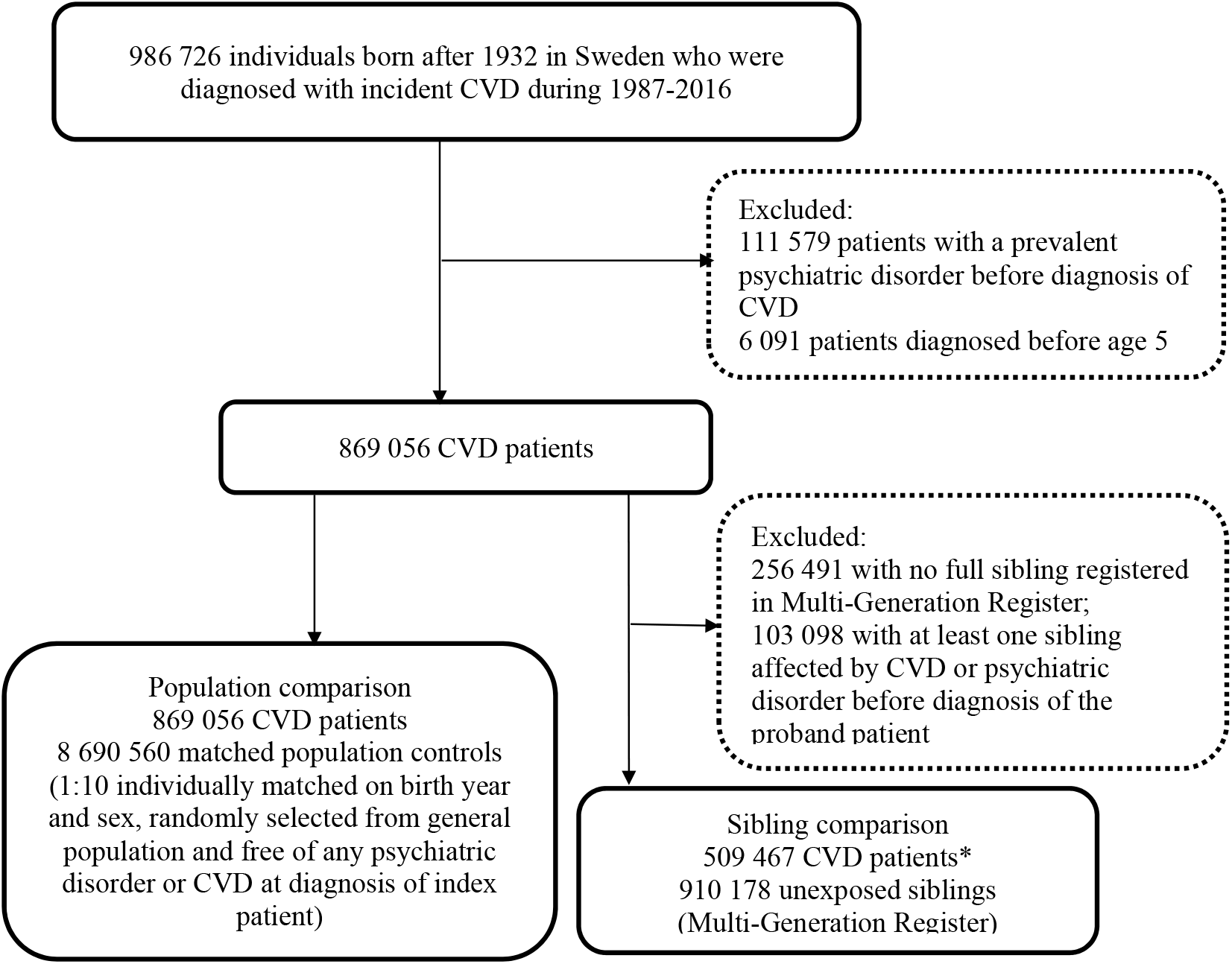
Study design. CVD: cardiovascular disease. * 67 745 families had more than one sibling affected by CVD.

**Table S2.** International Classification of Diseases (ICD) codes for exposure, outcome and covariates identifications.

|  | ICD-9 | ICD-10 |
| --- | --- | --- |
| **Any psychiatric disorder** | 291,292, 295-311 | F10-F69 |
| Non-affective psychotic disorders | 295, 297, 298, excluding 295H and 298B | F20-24, F28-29 |
| Affective psychotic disorders | 296, 295H, 298B | F25, F30-31, F32.3, F33.3 |
| Alcohol or drug misuse | 291, 292, 303, 304 | F10-16, F18-19 |
| Mood disorders, excluding those with psychotic symptoms | 300E, 311 | F32-34, F38-39, excluding F32.3 and F33.3 |
| Anxiety and stress related disorders | 300A-D, 300F-H, 300W-X, 306, 307A, 308, 309 | F40-48 |
| -Acute stress reaction | 308, 309A | F43.0 |
| Eating disorders | 307B, 307F | F50 |
| Personality disorders | 301 | F60-63, F68-69 |
| **Any cardiovascular disease** | 390-438, 440,444,445 | I00-I70, I730, I74- I75 |
| Ischemic heart disease | 410-414 | I20-I24, I25 (excluding I25.5) |
| Cerebrovascular disease | 430-434, 436-438 | I60-I69 |
| Emboli and thrombosis | 415, 444,445 | I26, I74, I75 |
| Hypertensive disease | 401-405 | I10-I16, I674 |
| Heart failure | 428 | I25.5, I42.0, I42.8, I42.9, I50 |
| Arrhythmia/conduction disorder | 426, 427 | I44-I49 |
| **Covariates: history of severe somatic conditions** |  |  |
| Chronic pulmonary disease | 500-505, 506C | J60-J67, J684, J70, J703 |
| Connective tissue disease | 710A, 710B, 710E, 714A, 714B, 714C, 714W,714X, 725 | M05, M06, M315, M32-M34, M351, M353, M360 |
| Diabetes | 250A, 250G | E100, E101, E106, E108, E109, E110, E111, E116, E118, E119, E120, E121, E126, E128, E129, E130, E131, E136, E138, E139, E140, E141, E146, E148, E149 |
| Diabetes with end organ damage | 250D-250F | E102-E105, E107, E112-E115, E117, E122-E125, E127, E132-E135, E137, E142-E145, E147 |
| Moderate or severe renal disease | 582, 583A, 583B, 583C, 583E, | I120, I131, N032-N037, N052-N057, N18, N19, N250, Z490-Z492, Z940, Z992 |
| Mild liver disease | 583G, 583H, 585, 586, 588 | B18, K700-K703, K709, K713-K715, K717, K73, K74, K760, K762--K764, K768, K769, Z944 |
| Moderate or severe liver disease | 571C, 571E,571F, 571G | I850, I859, I864, I982, K704, K711, K721, K729, K765, K766, K767 |
| Ulcer disease | 572C, 572D, 572E, 572W, 456A, 456B, 456C | K25-K28 |
| Any malignancy, including leukemia and lymphoma | 140-172, 174-195, 200-208 | C00-C26, C30-C34, C37-C41, C43, C45-C58, C60-C76, C81-C85, C88, C90-C97 |
| Metastatic cancer | 196,197,198,199A, 199B | C77- C80 |
| HIV/AIDS | 042, 043, 044 | B20-B22, B24 |

**Table S3.**
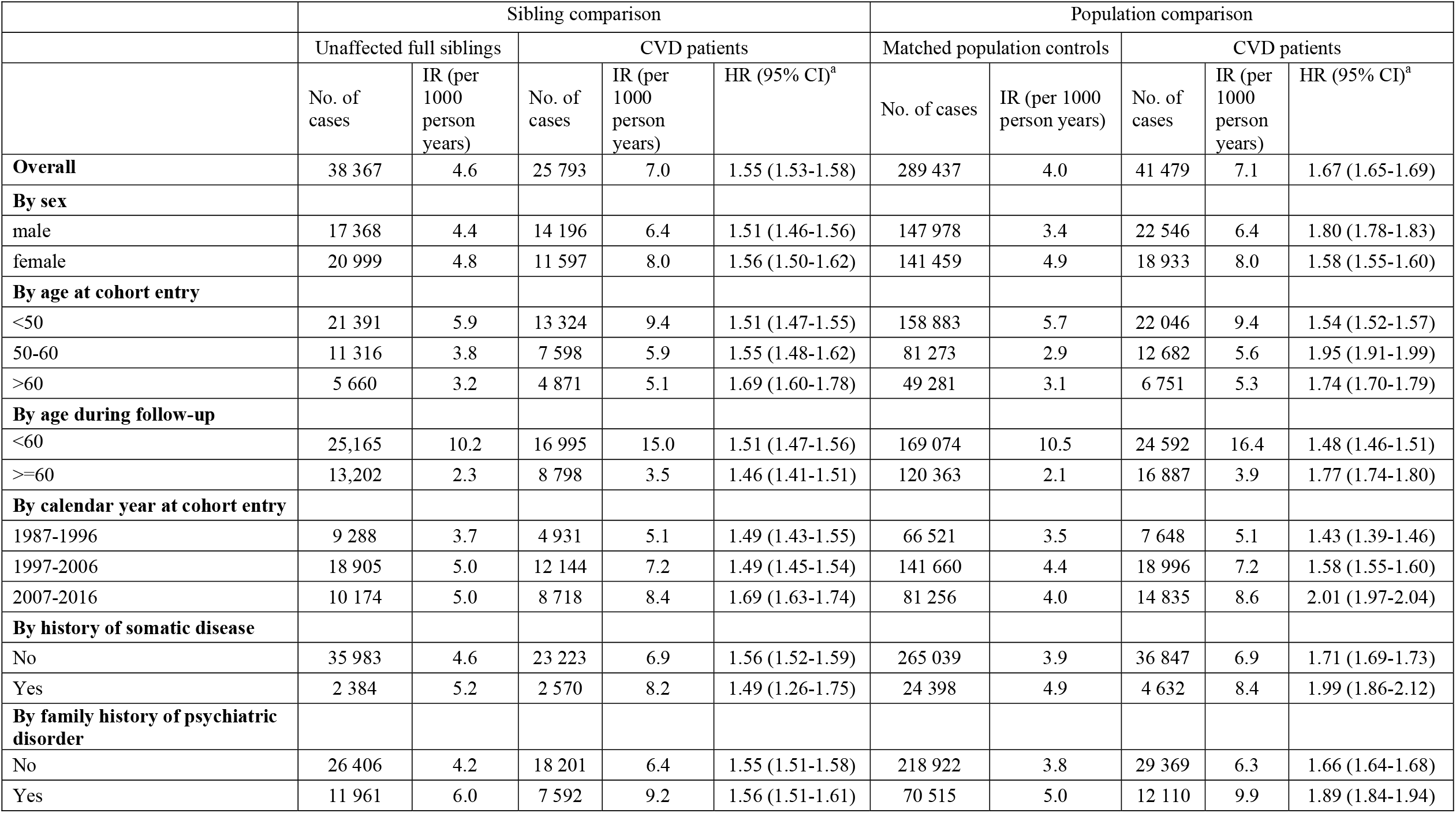
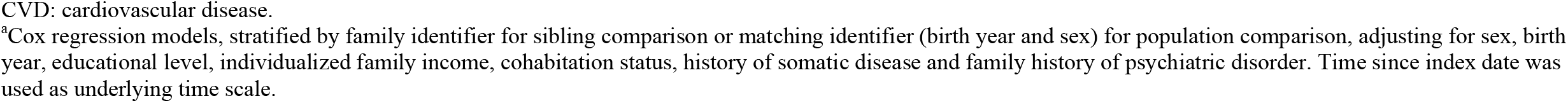
Crude incidence rates (IRs) and hazard ratios (HRs) with 95% confidence intervals (CIs) for incident psychiatric disorder among CVD patients compared with their full siblings or matched population controls, by patient characteristics.

|  | Sibling comparison |  |  |  |  | Population comparison |  |  |  |  |
| --- | --- | --- | --- | --- | --- | --- | --- | --- | --- | --- |
|  | Unaffected full siblings |  | CVD patients |  |  | Matched population controls |  | CVD patients |  |  |
|  | No. of cases | IR (per 1000 person years) | No. of cases | IR (per 1000 person years) | HR (95% CI)<sup>a</sup> | No. of cases | IR (per 1000 person years) | No. of cases | IR (per 1000 person years) | HR (95% CI)<sup>a</sup> |
| **Overall** | 38 367 | 4.6 | 25 793 | 7.0 | 1.55 (1.53-1.58) | 289 437 | 4.0 | 41 479 | 7.1 | 1.67 (1.65-1.69) |
| **By sex** |  |  |  |  |  |  |  |  |  |  |
| male | 17 368 | 4.4 | 14 196 | 6.4 | 1.51 (1.46-1.56) | 147 978 | 3.4 | 22 546 | 6.4 | 1.80 (1.78-1.83) |
| female | 20 999 | 4.8 | 11 597 | 8.0 | 1.56 (1.50-1.62) | 141 459 | 4.9 | 18 933 | 8.0 | 1.58 (1.55-1.60) |
| **By age at cohort entry** |  |  |  |  |  |  |  |  |  |  |
| <50 | 21 391 | 5.9 | 13 324 | 9.4 | 1.51 (1.47-1.55) | 158 883 | 5.7 | 22 046 | 9.4 | 1.54 (1.52-1.57) |
| 50-60 | 11 316 | 3.8 | 7 598 | 5.9 | 1.55 (1.48-1.62) | 81 273 | 2.9 | 12 682 | 5.6 | 1.95 (1.91-1.99) |
| >60 | 5 660 | 3.2 | 4 871 | 5.1 | 1.69 (1.60-1.78) | 49 281 | 3.1 | 6 751 | 5.3 | 1.74 (1.70-1.79) |
| **By age during follow-up** |  |  |  |  |  |  |  |  |  |  |
| <60 | 25,165 | 10.2 | 16 995 | 15.0 | 1.51 (1.47-1.56) | 169 074 | 10.5 | 24 592 | 16.4 | 1.48 (1.46-1.51) |
| >=60 | 13,202 | 2.3 | 8 798 | 3.5 | 1.46 (1.41-1.51) | 120 363 | 2.1 | 16 887 | 3.9 | 1.77 (1.74-1.80) |
| **By calendar year at cohort entry** |  |  |  |  |  |  |  |  |  |  |
| 1987-1996 | 9 288 | 3.7 | 4 931 | 5.1 | 1.49 (1.43-1.55) | 66 521 | 3.5 | 7 648 | 5.1 | 1.43 (1.39-1.46) |
| 1997-2006 | 18 905 | 5.0 | 12 144 | 7.2 | 1.49 (1.45-1.54) | 141 660 | 4.4 | 18 996 | 7.2 | 1.58 (1.55-1.60) |
| 2007-2016 | 10 174 | 5.0 | 8 718 | 8.4 | 1.69 (1.63-1.74) | 81 256 | 4.0 | 14 835 | 8.6 | 2.01 (1.97-2.04) |
| **By history of somatic disease** |  |  |  |  |  |  |  |  |  |  |
| No | 35 983 | 4.6 | 23 223 | 6.9 | 1.56 (1.52-1.59) | 265 039 | 3.9 | 36 847 | 6.9 | 1.71 (1.69-1.73) |
| Yes | 2 384 | 5.2 | 2 570 | 8.2 | 1.49 (1.26-1.75) | 24 398 | 4.9 | 4 632 | 8.4 | 1.99 (1.86-2.12) |
| **By family history of psychiatric disorder** |  |  |  |  |  |  |  |  |  |  |
| No | 26 406 | 4.2 | 18 201 | 6.4 | 1.55 (1.51-1.58) | 218 922 | 3.8 | 29 369 | 6.3 | 1.66 (1.64-1.68) |
| Yes | 11 961 | 6.0 | 7 592 | 9.2 | 1.56 (1.51-1.61) | 70 515 | 5.0 | 12 110 | 9.9 | 1.89 (1.84-1.94) |

CVD: cardiovascular disease.
<sup>a</sup>Cox regression models, stratified by family identifier for sibling comparison or matching identifier (birth year and sex) for population comparison, adjusting for sex, birth year, educational level, individualized family income, cohabitation status, history of somatic disease and family history of psychiatric disorder. Time since index date was used as underlying time scale.

**Table S4.** Crude incidence rates (IRs) and hazard ratios (HRs) with 95% confidence intervals (CIs) for incident psychiatric disorders among CVD patients compared with their full siblings or matched population controls, by time of follow-up (<1 or >=1 year from CVD diagnosis)

|  | Sibling comparison |  |  | Population comparison |  |  |
| --- | --- | --- | --- | --- | --- | --- |
|  | No. of cases | IR (per 1000 person years) | HR (95% CI)<sup>a</sup> | No. of cases | IR (per 1000 person years) | HR (95% CI)<sup>a</sup> |
| **<1 year follow-up** |  |  |  |  |  |  |
| Full siblings/matched population controls | 4 024 | 4.6 | 1.0 | 33 625 | 4.1 | 1.0 |
| CVD patients_controlled for sex, birth year, educational level, individualized family income and cohabitation status | 5 695 | 12.6 | 2.76 (2.64-2.89) | 9 545 | 12.6 | 3.06 (2.99-3.14) |
| As above + history of somatic diseases |  |  | 2.74 (2.62-2.87) |  |  | 3.02 (2.95-3.09) |
| As above + family history of psychiatric disorder |  |  | - |  |  | 3.00 (2.93-3.07) |
| **≥1 year follow-up** |  |  |  |  |  |  |
| Full siblings/ matched population controls | 36 021 | 4,8 | 1.0 | 271 900 | 4.3 | 1.0 |
| CVD patients_controlled for sex, birth year, educational level, individualized family income and cohabitation status | 23 955 | 6.9 | 1.46 (1.43-1.49) | 38 414 | 6.9 | 1.60 (1.58-1.61) |
| As above + history of somatic diseases |  |  | 1.45 (1.42-1.48) |  |  | 1.56 (1.54-1.57) |
| As above + family history of psychiatric disorder |  |  | - |  |  | 1.54 (1.53-1.56) |

CVD: cardiovascular disease.
<sup>a</sup>Cox regression models, stratified by family identifier for sibling comparison or matching identifier (birth year and sex) for population comparison. Time since index date was used as underlying time scale.

**Figure S2.**
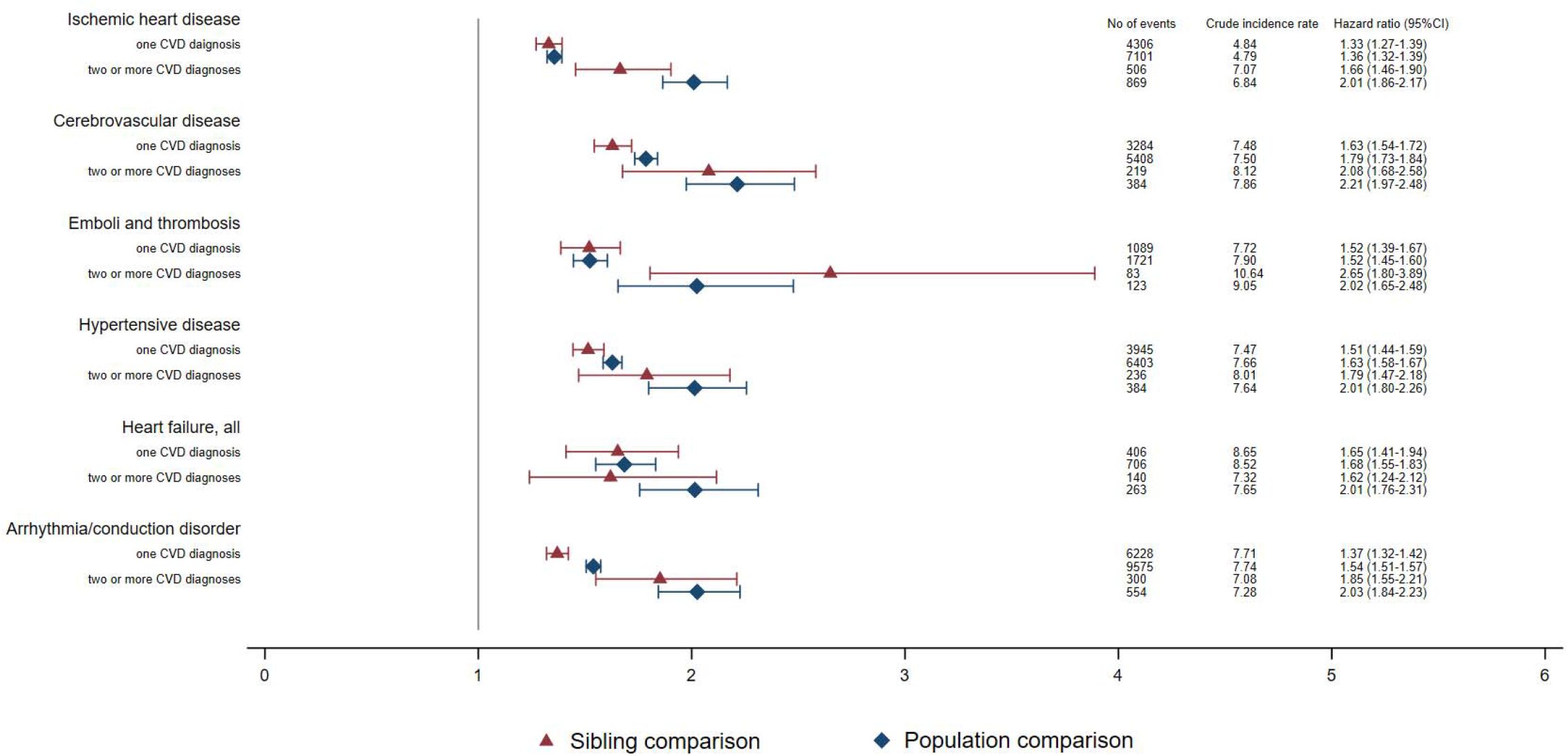
Crude incidence rates and hazard ratios with 95% confidence intervals (CIs) for an incident psychiatric disorder among different types of CVD patients compared with their full siblings or matched population controls, by number of CVD diagnoses during >=1 year of follow-up^a^. CVD: cardiovascular disease. ^a^ Cox regression models, stratified by family identifier for sibling comparison or matching identifier (birth year and sex) for population comparison, and controlled for educational level, individualized family income, cohabitation status, as well as sex and birth year (in sibling comparison). Time since index date was used as underlying time scale. For patients with two or more CVD diagnoses, follow up time started from diagnosis of that cardiovascular comorbidity.

**Table S5.** Crude incidence rates (IRs) and hazard ratios (HRs) with 95% confidence intervals (CIs) for psychiatric disorders among CVD patients compared with their full siblings or matched population controls, excluding CVD patients medicated with psychotropic drugs, by time of follow-up (<1 or >=1 year from CVD diagnosis)^a^.

|  | Sibling comparison<sup>§</sup> |  |  | Population comparison<sup>§</sup> |  |  |
| --- | --- | --- | --- | --- | --- | --- |
|  | No. of cases | IR (per 1000 person years) | HR (95% CI)<sup>b</sup> | No. of cases | IR (per 1000 person years) | HR (95% CI)<sup>b</sup> |
| **<1 year follow up** |  |  |  |  |  |  |
| Full siblings/matched population controls | 12 439 | 35.4 | 1.0 | 129 332 | 36.3 | 1.0 |
| CVD patients_controlled for sex, birth year, educational level, individualized family income and cohabitation status | 22 609 | 122.2 | 3.73 (3.62-3.83) | 40 008 | 130.5 | 3.62 (3.57-3.66) |
| As above + history of somatic diseases |  |  | 3.70 (3.60-3.81) |  |  | 3.60 (3.56-3.65) |
| As above + family history of psychiatric disorder |  |  | - |  |  | 3.61 (3.56-3.65) |
| **≥1 year follow up** |  |  |  |  |  |  |
| Full siblings/ matched population controls | 45 253 | 32.2 | 1.0 | 466 103 | 33.8 | 1.0 |
| CVD patients_controlled for sex, birth year, educational level, individualized family income and cohabitation status | 31 508 | 43.6 | 1.41 (1.39-1.44) | 55 074 | 47.4 | 1.39 (1.37-1.40) |
| As above + history of somatic diseases |  |  | 1.40 (1.38-1.43) |  |  | 1.39 (1.38-1.41) |
| As above + family history of psychiatric disorder |  |  | - |  |  | 1.40 (1.38-1.41) |

CVD: cardiovascular disease.
<sup>a</sup> CVD patients diagnosed during 2006-2016 were included in this analysis due to the availability of data on prescribed drug. In sibling comparison, 27.8% CVD patients and 23.5% siblings were excluded due to prior medication of psychotropic drugs before index date. In population comparison, 30.6% CVD patients and 25.0% population controls were excluded due to prior medicated with psychotropic drugs before index date.
<sup>b</sup> Cox regression models, stratified by family identifier for sibling comparison or matching identifier (birth year and sex) for population comparison. Time since index date was used as underlying time scale. Definition of psychiatric disorder included hospital visits as well as use of psychotropic drugs during follow-up.

**Table S6.**
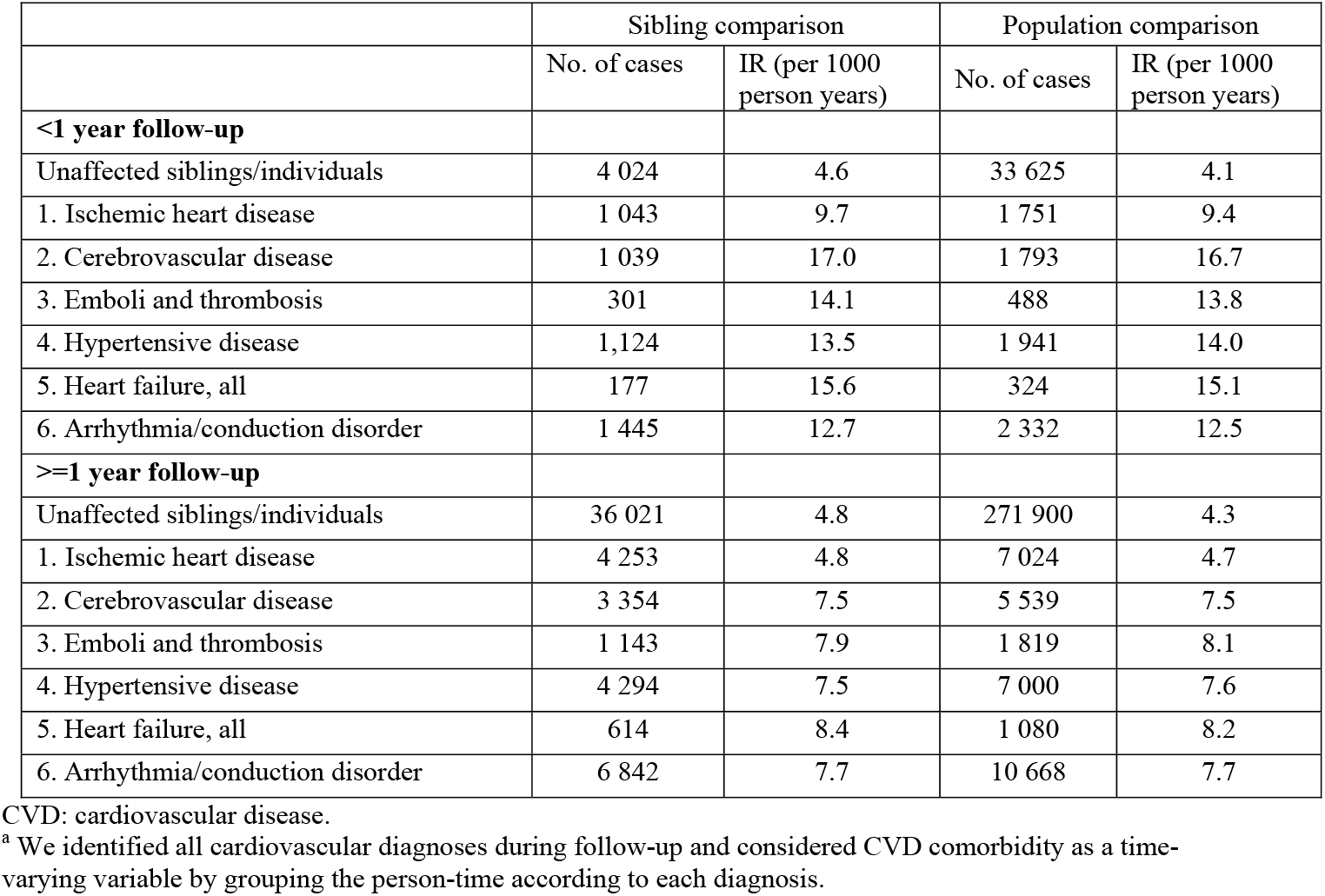
Crude incidence rates (IRs) for psychiatric disorders among different groups of CVD patients, their full siblings and matched population controls, by time of follow-up (<1 or >=1 year from CVD diagnosis)^a^.

|  | Sibling comparison |  | Population comparison |  |
| --- | --- | --- | --- | --- |
|  | No. of cases | IR (per 1000 person years) | No. of cases | IR (per 1000 person years) |
| **<1 year follow-up** |  |  |  |  |
| Unaffected siblings/individuals | 4 024 | 4.6 | 33 625 | 4.1 |
| 1. Ischemic heart disease | 1 043 | 9.7 | 1 751 | 9.4 |
| 2. Cerebrovascular disease | 1 039 | 17.0 | 1 793 | 16.7 |
| 3. Emboli and thrombosis | 301 | 14.1 | 488 | 13.8 |
| 4. Hypertensive disease | 1,124 | 13.5 | 1 941 | 14.0 |
| 5. Heart failure, all | 177 | 15.6 | 324 | 15.1 |
| 6. Arrhythmia/conduction disorder | 1 445 | 12.7 | 2 332 | 12.5 |
| **≥1 year follow-up** |  |  |  |  |
| Unaffected siblings/individuals | 36 021 | 4.8 | 271 900 | 4.3 |
| 1. Ischemic heart disease | 4 253 | 4.8 | 7 024 | 4.7 |
| 2. Cerebrovascular disease | 3 354 | 7.5 | 5 539 | 7.5 |
| 3. Emboli and thrombosis | 1 143 | 7.9 | 1 819 | 8.1 |
| 4. Hypertensive disease | 4 294 | 7.5 | 7 000 | 7.6 |
| 5. Heart failure, all | 614 | 8.4 | 1 080 | 8.2 |
| 6. Arrhythmia/conduction disorder | 6 842 | 7.7 | 10 668 | 7.7 |

CVD: cardiovascular disease.
<sup>a</sup> We identified all cardiovascular diagnoses during follow-up and considered CVD comorbidity as a time-varying variable by grouping the person-time according to each diagnosis.

**Table S7.**
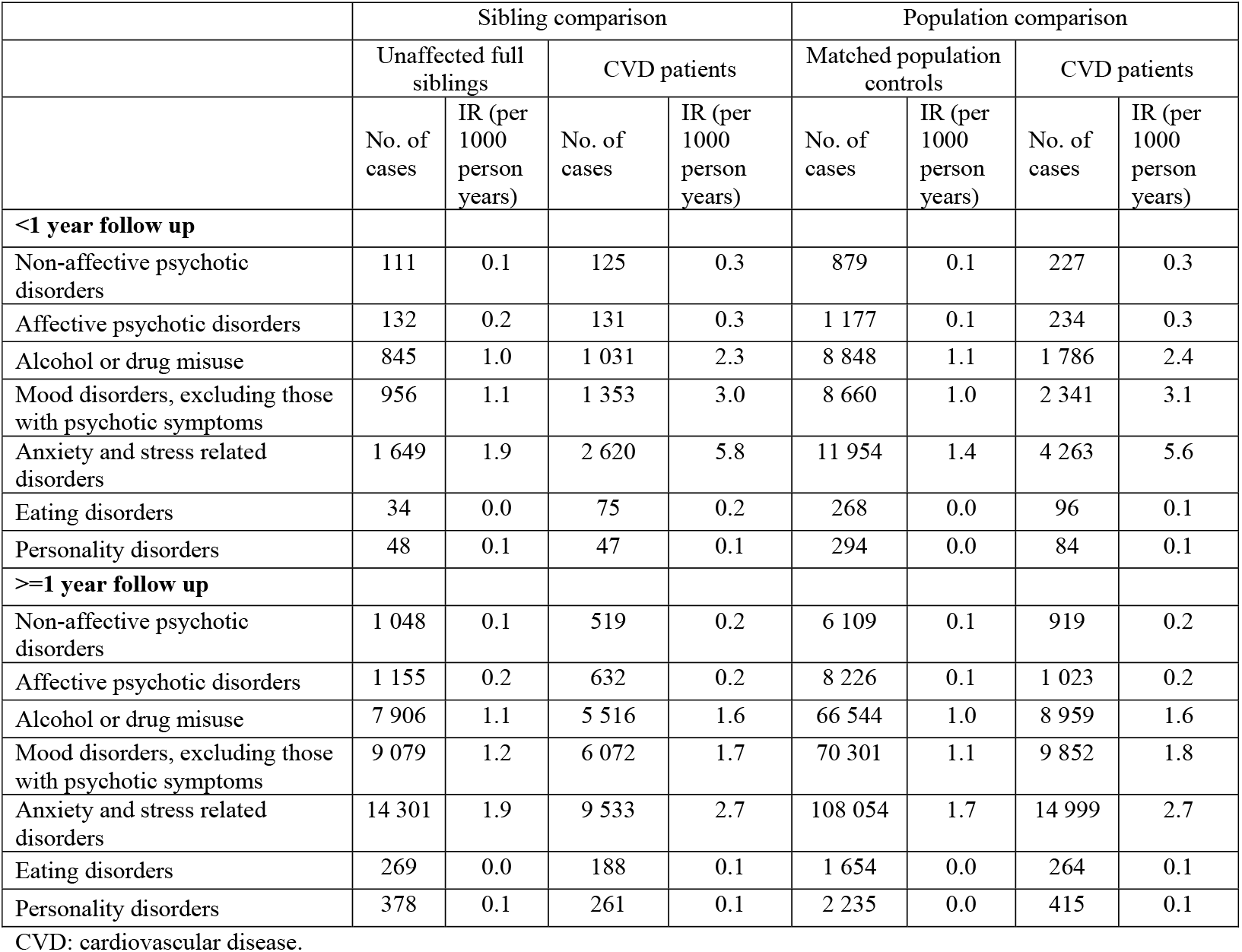
Crude incidence rates (IRs) of different types of psychiatric disorder among CVD patients, their full siblings, and matched population controls, by time of follow-up (<1 or >=1 year from CVD diagnosis)

|  | Sibling comparison |  |  |  | Population comparison |  |  |  |
| --- | --- | --- | --- | --- | --- | --- | --- | --- |
|  | Unaffected full siblings |  | CVD patients |  | Matched population controls |  | CVD patients |  |
|  | No. of cases | IR (per 1000 person years) | No. of cases | IR (per 1000 person years) | No. of cases | IR (per 1000 person years) | No. of cases | IR (per 1000 person years) |
| **<1 year follow up** |  |  |  |  |  |  |  |  |
| Non-affective psychotic disorders | 111 | 0.1 | 125 | 0.3 | 879 | 0.1 | 227 | 0.3 |
| Affective psychotic disorders | 132 | 0.2 | 131 | 0.3 | 1 177 | 0.1 | 234 | 0.3 |
| Alcohol or drug misuse | 845 | 1.0 | 1 031 | 2.3 | 8 848 | 1.1 | 1 786 | 2.4 |
| Mood disorders, excluding those with psychotic symptoms | 956 | 1.1 | 1 353 | 3.0 | 8 660 | 1.0 | 2 341 | 3.1 |
| Anxiety and stress related disorders | 1 649 | 1.9 | 2 620 | 5.8 | 11 954 | 1.4 | 4 263 | 5.6 |
| Eating disorders | 34 | 0.0 | 75 | 0.2 | 268 | 0.0 | 96 | 0.1 |
| Personality disorders | 48 | 0.1 | 47 | 0.1 | 294 | 0.0 | 84 | 0.1 |
| **≥1 year follow up** |  |  |  |  |  |  |  |  |
| Non-affective psychotic disorders | 1 048 | 0.1 | 519 | 0.2 | 6 109 | 0.1 | 919 | 0.2 |
| Affective psychotic disorders | 1 155 | 0.2 | 632 | 0.2 | 8 226 | 0.1 | 1 023 | 0.2 |
| Alcohol or drug misuse | 7 906 | 1.1 | 5 516 | 1.6 | 66 544 | 1.0 | 8 959 | 1.6 |
| Mood disorders, excluding those with psychotic symptoms | 9 079 | 1.2 | 6 072 | 1.7 | 70 301 | 1.1 | 9 852 | 1.8 |
| Anxiety and stress related disorders | 14 301 | 1.9 | 9 533 | 2.7 | 108 054 | 1.7 | 14 999 | 2.7 |
| Eating disorders | 269 | 0.0 | 188 | 0.1 | 1 654 | 0.0 | 264 | 0.1 |
| Personality disorders | 378 | 0.1 | 261 | 0.1 | 2 235 | 0.0 | 415 | 0.1 |

CVD: cardiovascular disease.

**Figure S3.**
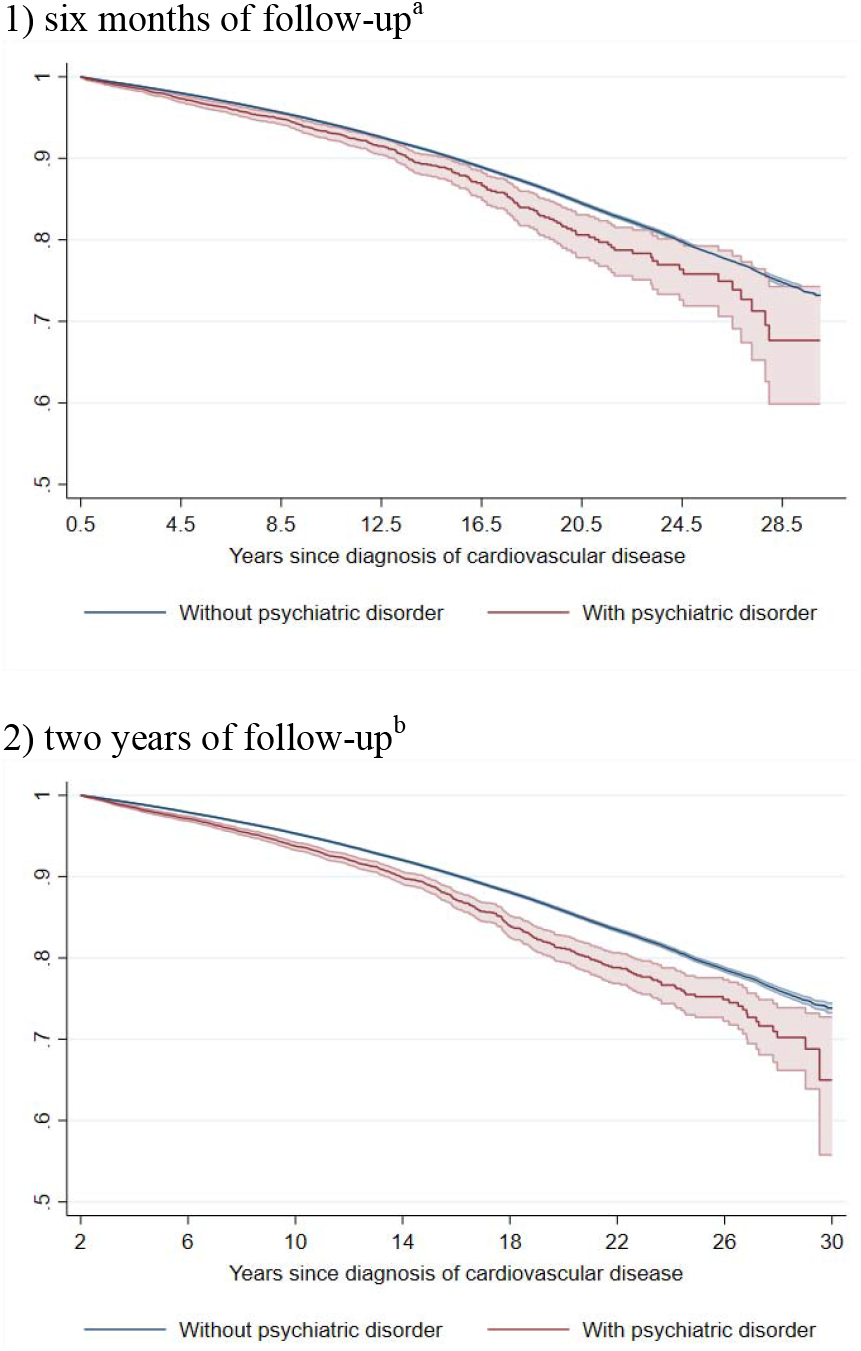
Kaplan-Meier estimates of CVD death in CVD patients with and without incident psychiatric disorder during 1) six months or 2) two years of follow-up. CVD: cardiovascular disease. ^a^ Time since index date was used as underlying time scale. 94.1% of CVD patients (N=817 748) survived the six months of follow-up and included in this analysis. The hazard ratio of cardiovascular death was 1.40 (95% confidence interval 1.27 to 1.54) when comparing CVD patients with psychiatric disorder to patients without such a psychiatric diagnosis (mortality rate, 8.1 and 7.0 per 1000 person-years, respectively). ^b^ Time since index date was used as underlying time scale. 83.8% of CVD patients (N=728 179) survived the two years of follow-up and included in this analysis. The hazard ratio of cardiovascular death was 1.52 (95% confidence interval 1.43 to 1.62) when comparing CVD patients with psychiatric disorder to patients without such a psychiatric diagnosis (mortality rate, 9.1 and 7.4 per 1000 person-years, respectively).

**Figure S4.**
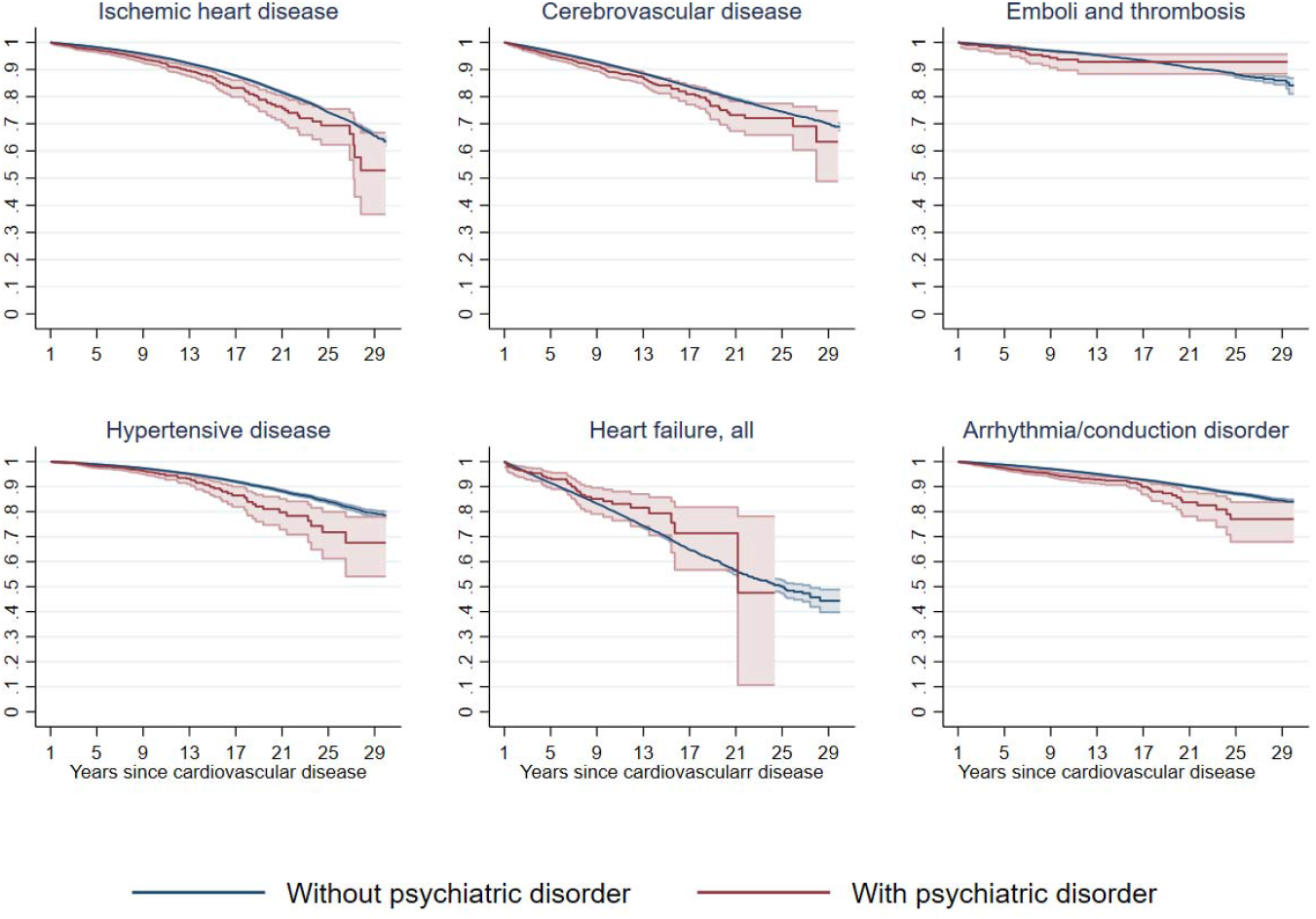
Kaplan-Meier estimates of CVD death in CVD patients with and without incident psychiatric disorder during the first year of follow-up, according to types of first CVD diagnosis^a^. CVD: cardiovascular disease. ^a^ Time since index date was used as underlying time scale. 90.4% of CVD patients (N=785 287) survived the first year of follow-up and included in this analysis.

